# Investigation of Measles Outbreak in Pakistan (2022 to 2023): Exploring Comorbidities, Complications, and Molecular Dynamics

**DOI:** 10.1101/2025.11.20.25340652

**Authors:** Asia Nawaz, Sidra Rahman, Nighat Haider, Adnan Zeb, Muhammad Ali

**Affiliations:** Department of Biotechnology, Quaid-i-Azam University Islamabad, Pakistan; Pediatric Infectious Diseases Unit, Children Hospital Pakistan Institute of Medical Sciences (PIMS), ICT Islamabad, Pakistan

**Keywords:** Measles, Epidemiology, Nucleoprotein gene, Phylogenetic Analysis, B3 genotype, Pakistan

## Abstract

**Background:** Measles remains a major public health concern worldwide, particularly in regions with insufficient vaccination coverage and healthcare infrastructure like Pakistan. The purpose of this study is to investigate that what are the epidemiological, molecular, and phylogenetic characteristics of the measles outbreak in Pakistan during 2022-2023 and highlight the areas of improvement that can contribute to the understanding of the disease and inform strategies for effective outbreak management and vaccination optimization.

**Methods:** Throat swabs were collected from patients (n = 183) admitted at PIMS, Islamabad, Pakistan from December 2022-December 2023. This research was conducted statistically by using SPSS-21 software. The phylogenetics, amino acid substitutions, and structural analysis of partial nucleoprotein were performed by using different bioinformatics software.

**Results:** Most of the measles patients were infants with a high prevalence of comorbidity and complications and significant patients had inadequate vaccination coverage. The phylogenetic analysis revealed relatedness to B3 strains circulating in Russia and the USA, emphasizing the global spread. The amino acid substitutions and structure analysis highlighted minor structural variations in current isolates as compared to the reference strain.

**Conclusion:** The findings underscore the importance of continuous surveillance and understanding of measles epidemiology and viral variations for effective outbreak management and vaccination optimization, ultimately aiming to mitigate the morbidity and mortality associated with measles outbreaks in Pakistan and similar settings globally. This study exemplifies the significance of interdisciplinary collaboration in confronting complex infectious disease challenges, emphasizing the imperative of continuous monitoring and preparedness in combating measles through vaccination.

## 1. Introduction

Measles continues to be a major contributor to the global disease burden. It is a highly infectious disease caused by the Measles virus that belongs to the genus *Morbillivirus* within the family *Paramyxoviridae*. The measles virus is serologically monotypic and has been genetically characterized into eight clades (A-H) and 24 genotypes (A, B1-3, C1-2 D1-11, E, F, G1-3, and H1-2). Molecular and epidemiological studies elucidate the spatiotemporal origin of circulating viruses and highlight any changes in the typical pattern and severity of disease in an endemic region [1]. Despite the availability of an effective measles vaccine for 50 years, it is still causing significant mortality and morbidities among young children in underdeveloped countries specifically targeting the younger population in resource-poor settings [2]. Measles clinically presents itself as fever cough, coryza, maculopapular rash, and conjunctivitis. In severe cases, it can lead to several neurological, gastrointestinal, and respiratory complications and even death [3]. Measles affects all the organs of the body via infecting mucous membranes and immunosuppression due to the prolonged infection renders the patient prone to several secondary infections. The mode of transmission for measles is through respiratory droplets and aerosols and having a high reproduction number (R0) around 12-18 in susceptible populations can be quite contagious [4]. The incubation period of measles is approximately two weeks, and the patient recovers within three weeks if the infection is mild. In other cases when the infection is persistent it can take serious form and lead to severe complications such as pneumonia that is fatal pertinent to 56-86% of measles-related deaths. In around 50% of pneumonia there is bacterial superinfection [5]. Diarrhea and brain infections including encephalitis, and meningitis are also major causes of death among measles patients. Encephalitis is caused by the primary viral infection if the virus makes its way into the brain of an infected person and is followed by chemokines and lymphokines infiltration. In this situation around 10-15% of patients succumb to death and if the virus persists it can lead to post-infection Encephalomyelitis (PIE) that causes permanent damage to the nervous system [6–7].

From 2000 to 2021, around 56 million deaths were averted due to extensive immunization in different measles-endemic regions. In 2021 a mass measles and rubella vaccination campaign was launched in Pakistan that aimed to immunize 90 million children between 9 months to 15 years [8]. As a result, the overall vaccine coverage remained at 81%. During the summer of 2022 Pakistan was hit by major floods that affected the health infrastructure and massive displacements into temporary settlements and camps provided breeding grounds for measle outbreaks that reared its head in 31 districts of Pakistan. According to data from the National Disaster Management Authority 12 out of 13 (almost 92%) of these measles-endemic areas were flood-affected. By the end of 2022 another measles immunization campaign was launched by EPI that targeted 1.8 million children from 38 districts to prevent the further escalation in measles cases and it covered 98% of the child population between 6-59 months successfully [9]. Despite all these efforts made by the local and international organizations measles is still looming around. A huge resurgence has been witnessed where the global measles cases increased by 55% from 171,156 to 306291 during 2022-2023 [10]. As per a surveillance report by the CDC during the first half of 2023, approximately 10,549 measles cases were reported from Pakistan, making it the third major contributor to the measles global burden following India and Yemen, and during July-December 2024 the number of cases are still significant at 7,148 which brings Pakistan on the top of the list for the countries constantly battling with the measles disease burden [11].

In 2022-2023, a substantial measles outbreak in Pakistan necessitated a multifaceted study that employs epidemiological and molecular methodologies to elucidate the outbreak’s complexities. This research explores comorbidities, complications, and molecular dynamics of the measles virus (MV), thereby advancing our comprehension of this critical public health issue. Moreover, the substitutional analysis of the circulating virus is an effort to dig deeper into the structural and functional variation in the measles virus and find out possible associations between the modifications in the virus and the severity of the disease during the current outbreak in Pakistan.

## 2. Methodology

### 2.1. Ethical Approval

The current research was approved by the Bio-Ethical Committee (BEC-FBS-QAU2023-520) at Quaid-i-Azam University Islamabad.

### 2.2. Sample population

Throat swabs were collected from patients (n = 183) admitted at Children’s Hospital measles ward, Pakistan Institute of Medical Sciences (PIMS) Islamabad, Pakistan during December 2022-December 2023. All subjects showed typical measles manifestations with symptoms such as fever, cough, rash, conjunctivitis, *etc*. The enrolled subjects were aged between 3 months and 9 years of age and were divided into four age groups i.e. infants (0-12 months), Toddlers (13-36 months), Early Childhood (37-72 months), and Middle childhood (73-144 months). All the biological samples were collected after the written consent from the guardian/parent of the patients. Patient’s history and data including clinical, epidemiological, and vaccination status, were recorded from their hospital admission forms. Throat swab samples were transported to the Infectious Diseases and Molecular Pathology Laboratory (IDMPL), Quaid-I-Azam University Islamabad.

### 2.3. Statistical Analysis

Statistical analysis of patient’s clinical data was performed using SPSS-21 software. The frequency of symptoms and complications presented by the patients and duration of stay at the hospital among different age groups and both genders were analyzed using descriptive statistics. The normality test was carried out followed by the Kruskal Wallis test for comorbidities *w.r.t*., age groups. A *p-*value below 0.05 was considered significant. Post-hoc Mann-Whitney U test was used for different age groups and genders *w.r.t.*, average stay at hospital and comorbidities.

### 2.4. Primer designing

The primers for the N gene were designed on the NCBI primer designing tool, primer blast (https://www.ncbi.nlm.nih.gov/tools/primer-blast/), by using reference sequence NC_001498.1. Forward primer 5’ATCCTGCTCTTGGACTGCAT’3 and reverse primer 5’TGGGAGTGGATGGTTGGTTG’3 was used for amplification of partial sequence of N gene including N-450 region that has been recommended by Global Measles and Rubella Laboratories Network (GMRLN) for Measles genotyping. The specifically designed primers anneal to a region spanning nucleotides 949-1794 in the MV genome, generating an amplicon of approximately 846 bp in size.

### 2.5. Viral RNA Extraction and synthesis of complementary DNA (cDNA) by reverse transcription

Viral RNA was extracted from throat swab samples of measles patients using WizPrep™ Total RNA Mini Kit by following the standard manufacturer’s protocol and the extraction was carried out in a designated extraction room with negative pressure and equipped with a biosafety cabinet Class II type B2. The complementary DNA was synthesized from extracted RNA using RevertAid first strand cDNA synthesis kit (Thermo Scientific, USA) and reverse primer that was designed for the N gene. The composition of the reaction mixture for reverse transcription of a total 20 μl reaction is given as follows; 5x reaction buffer 4 μl, 10mM dNTP 2 μl, specific primer 2 μl, Revert Aid transcriptase 1 μl, nuclease-free water 2.5 μl, RiboLock (RNase inhibitor) 0.5 μl, Template RNA 8 μl. Reaction mixture incubation was performed at 25 °C for 5 minutes and 42 °C for 60 min, followed by 45 °C for 30 min and a heat inactivation step (70 °C for 5 min).

### 2.6. PCR amplification of N gene (partial)

The amplification was carried out by using specifically designed and optimized primers for the conserved region of the measles N gene. The 20 μl of PCR mixture contained 10 μl HI Fidelity Phusion master mix (Thermo Scientific, USA) forward and reverse primers (2 μl), template cDNA (2 μl), and distilled water 6 μl. The cyclic condition of PCR was 5 minutes at 96°C, followed by 35 cycles with 1 cycle comprising 45 s at 96 °C, 30 s at 58 °C and 1 min at 72 °C with and final extension of 72°C for 10 min.

### 2.7. Visualization by gel electrophoresis and nucleotide sequencing

The amplified PCR products were run on agarose gel (2%) followed by visualization under UV trans illuminator. The targeted bands were excised from the gel and purified using the GeneJET Gel Extraction Kit (Thermo-Scientific, USA) following the standard protocol provided by the manufacturer. The eluted product (amplified product of N gene partial sequence) was stored at -20°C. The PCR product was sequenced via Sanger sequencing with the gene-specific forward primers. The deduced sequences obtained by Sanger sequencing were visualized by using the BioEdit software package [12].

### 2.8. Alignment and phylogenetic analysis of nucleotide sequences

The analysis of the obtained sequences was carried out utilizing genetic data resources for the measles virus N gene that were available at the National Center for Biotechnology Information (NCBI). Retrieved sequences were aligned at BioEdit, and MEGA-11 software was used for the construction of a phylogenetic tree [13] by using the Neighbors joining method, and the bootstrap values were determined by 1000 replicates in MEGA-11 [14]. The sequencing data for all the viral isolates were submitted to GenBank.

### 2.9. Alignment and analysis of amino acid sequences

The nucleotide sequences were translated into amino acid sequences using the Expasy tool. The amino acid sequences were also aligned on MEGA-11 software, and then aligned via CLC sequence viewer (version 8) software with a reference sequence of WLY62623.1 reported from Russia.

### 2.10. Prediction of Secondary and Tertiary Structures

The secondary structure of all isolates and reference proteins was predicted through the online server PSIPRED (http://bioinf.cs.ucl.ac.uk/psipred/) (Accessed on 03, April 2024). The 2D Model of amino acid sequences was generated through the server by using PSI-BLAST and neural networking [15]. The tertiary structure of isolates was predicted through the Robetta server (https://robetta.bakerlab.org/) (Accessed on 03 April 2024). To construct the 3D structure of the protein, the server searches for an appropriate amino acid sequence template which is used for comparison modeling. If the amino acid of the template is missing the server utilizes de novo Rosetta fragment insertion approach [16]. For the prediction of 3D structures and identification of amino acid substitutions, Phymol software was utilized.

## 3. Results

In this study we investigated the population characteristics of 183 measles patients <9 years admitted to children’s hospital at Pakistan Institute of Medical Sciences (PIMS), Islamabad from December 2022 to December 2023, revealing notable demographic trends and comorbidity prevalence among the children of different age groups.

### 3.1. Population characteristics

Data of total 183 patients showed the mean age of the measles patients included in this study was 16.75 ± 1.6 months with a majority being male 52.45% and infants including children <12 months comprising 73.77% of total admitted patients. Patients (13.66%) displayed a variety of comorbidities including congenital, neurological, and respiratory complications at the time of hospital admission (Table. 1). A large fraction of patients i.e., 96.73% were discharged from the hospital after an average duration of 5.74 ± 0.2 days of hospitalization and few were expired (2.18%) because of complications. Patients (60.10%) were given antiviral therapy in the form of oral ribavirin as a part of their treatment. Among all the patients a majority (44.26%) had not received any dose of measles vaccine followed by partially vaccinated (28.96%) having their 1^st^ dose of measles vaccine and fully vaccinated (26.77%) with 2^nd^ dose of measles vaccine. There was no significant difference (*p* = 0.953) between the average duration of hospitalization between different age groups.

### 3.2 Incidence of complications

The results showed a high incidence of complications among infants *w.r.t*., to age groups and among male measles patients. Rash, pneumonia, and conjunctivitis were the most prevalent complications followed by gastrointestinal (GI) symptoms, seizures, encephalitis, croup, and hepatitis (Fig.1).

**Fig 1.**
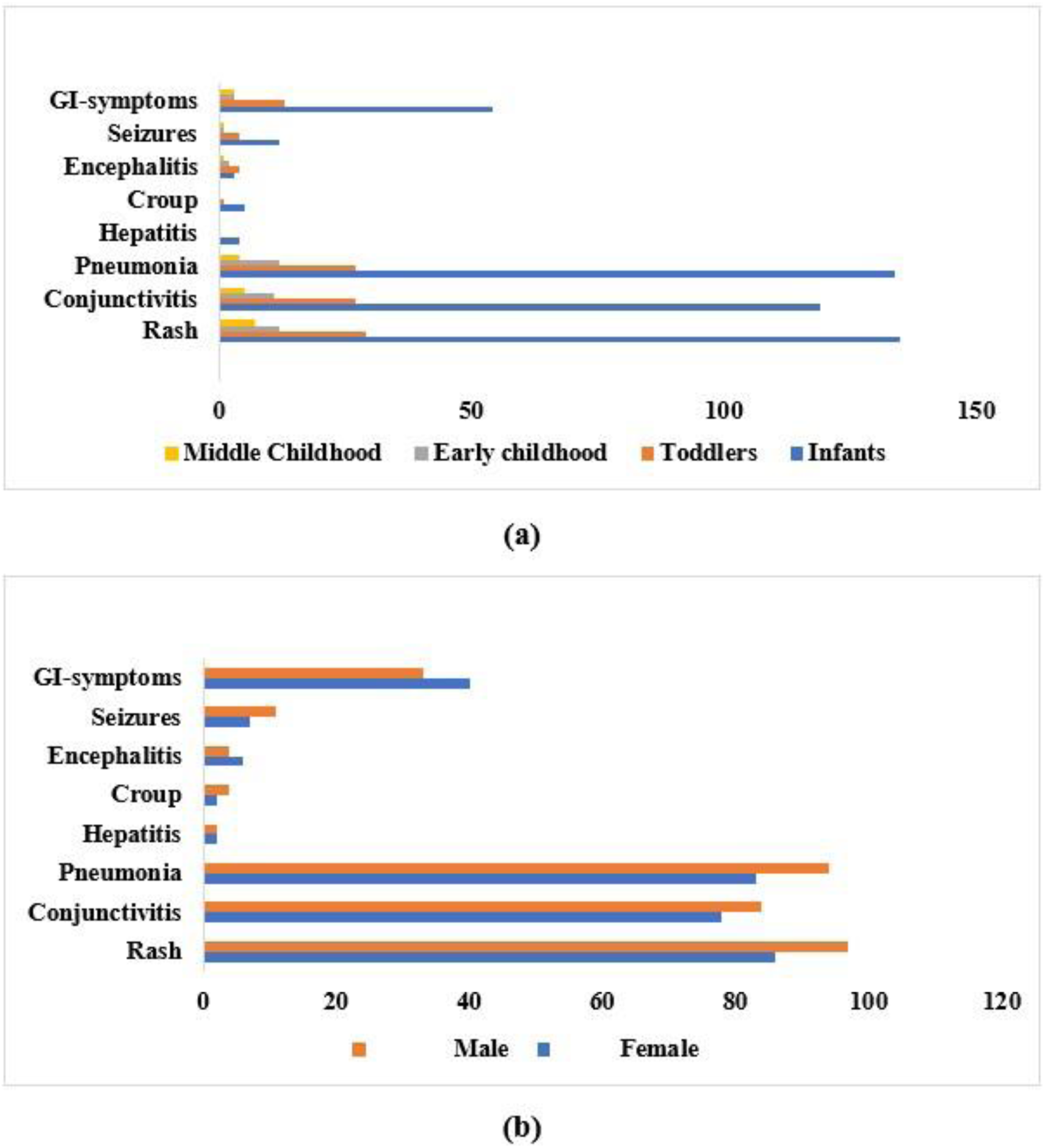
The incidence of complications in measles patients. (a) Presence of complications *w.r.t.,* different age groups (b) frequency of complications *w.r.t.,* gender

### 3.3. Results of statistical analysis

The duration of hospital stays varied significantly among different age groups. Specifically, the stay was significantly longer for infants (*p* = 0.000) and Toddlers (*p =* 0.000), but there was no significant difference between these two groups (*p =* 0.953). In contrast, the duration of hospital stay did not differ significantly between genders (*p =* 0.451).

Regarding the association of symptoms with different age groups, most of the symptoms including conjunctivitis, hepatitis, croup, seizures, and gastrointestinal symptoms showed no significant difference (*p*>0.05). However, except pneumonia (*p =* 0.000) and encephalitis (*p =* 0.014) were significantly different among different age groups. Further group-wise comparison revealed that pneumonia was significantly different between, Infants-Toddlers (*p =* 0.025), Infants-Middle childhood (*p =* 0.000), Toddlers-Middle Childhood (*p =* 0.015), Early Childhood-Middle Childhood (*p =* 0.016). Similarly, Encephalitis was significantly different between Infants-Toddlers (*p =* 0.005), and Infants-Early Childhood (*p =* 0.008). Lastly, the analysis of symptoms with respect to gender showed no significant differences (*p>*0.05), indicating that symptoms prevailed equally among both genders.

### 3.4. Sequencing of positive measles virus sample

The amplified product was purified from gel and subjected to Sanger sequencing. The obtained sequences of MV N-gene were further processed and analyzed by using different bioinformatics tools.

### 3.5. Phylogenetic analysis of partial nucleoprotein gene sequence of MVs

To evaluate the evolutionary history of our viral isolates we carried out phylogenetic analysis with the other closely related previously reported sequences by using the Neighbor-Joining method. The data for the previously reported measles virus isolates was retrieved from GenBank. The results of the evolutionary analysis revealed that our sequenced samples *(QAUPAK1, QAUPAK2, QAUPAK3, QAUPAK4 isolates*) were closely related to the measles virus reported from Russia and USA (accession number: OR290098 and OP314496) (Fig. 2). The sequences obtained from the viral isolates used in our current research were submitted to GenBank under the accession number OR622937, PP502160, PP502161, and PP502162.

**Fig 2.**
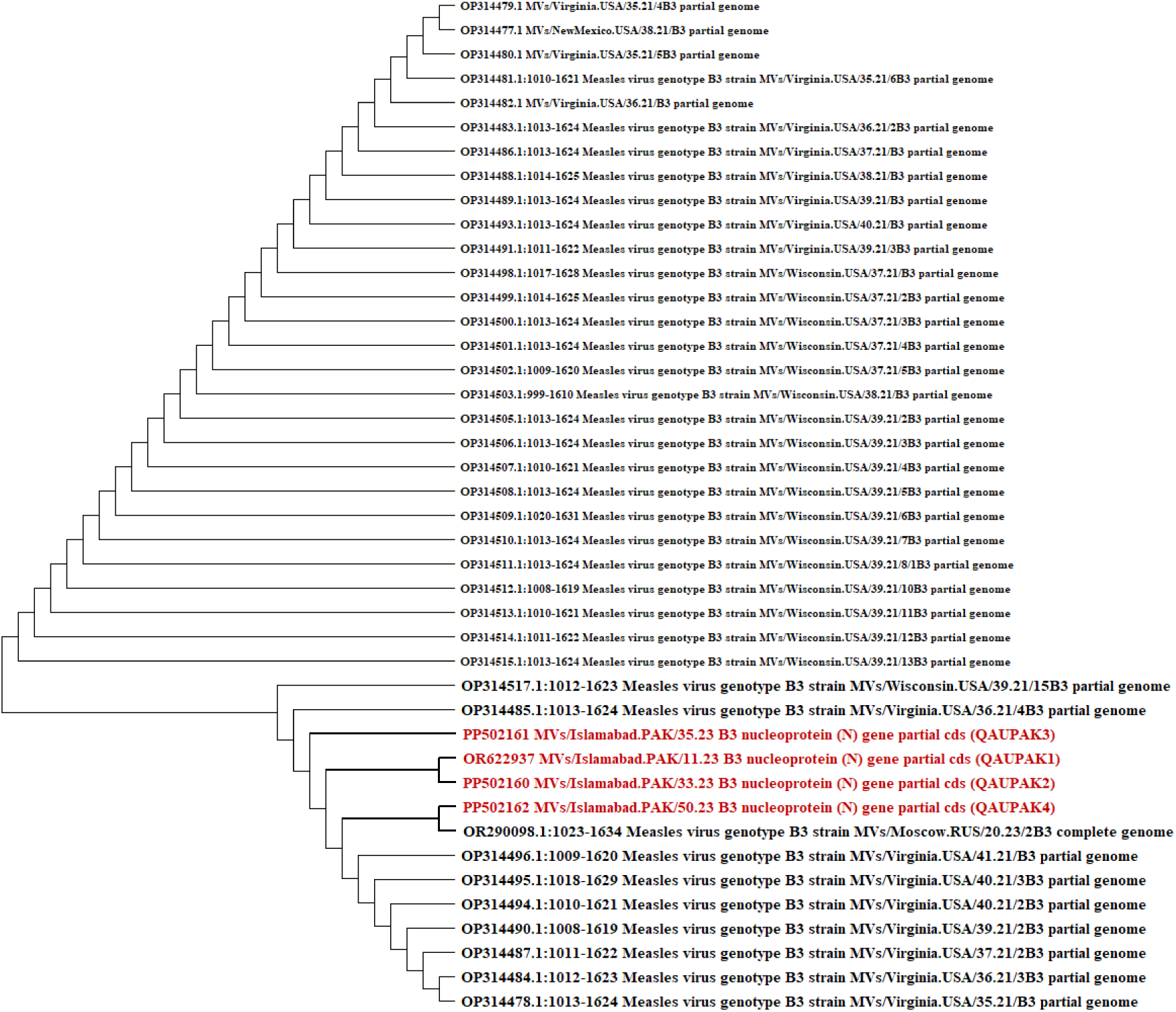
Evolutionary analyses of measles virus from Pakistan (n = 4) with the reference sequence reported globally (n = 38). The viral isolate in the current study clustered together and are closely related to viral isolates reported from Russia and USA which shows that they are phylogenetically related

### 3.7. Substitutional analysis in amino acid sequence

We translated our nucleotide sequence into amino acid sequences using Expasy translate tool and aligned them with the CLC sequence viewer. Amino acid sequences (Isolate names: QAUPAK1 QAUPAK2, QAUPAK3, QAUPAK4) were aligned with reference sequence WLY62623 reported from Russia due to phylogenetic proximity. The alignment of our sequences did not show any abnormalities in the sequence such as gaps, insertions, or deletions, *etc*. The analysis of amino acid revealed close similarities with the measles virus sequences that had been reported from Russia previously. There were a total of 3 substitutions, identified by the amino acid sequence analysis of query sequences *w.r.t.*, reference sequence. According to the reference sequence, QAUPAK1 isolate showed substitution at V394D, whereas S422G substitution was uniform for QAUPAK1, QAUPAK2, and QAUPAK3. Another substitution S487R was recorded in QAUPAK4. The details of substitutions have been presented in Table 2 and Fig 3.

**Table 1.** Overview of the characteristics of the study population

| Sr No. | Variables | Incidence in the study population |
| --- | --- | --- |
| i. | **Age** |  |
|  | i. Infant (0-12m) | 135 (73.77%) |
|  | ii. Toddlers (13-36 m) | 29 (15.84%) |
|  | iii. Early Childhood (37-72m) | 12 (6.55%) |
|  | iv. Middle Childhood (72-144m) | 7 (3.82%) |
| ii. | **Sex** |  |
|  | i. Male | 96 (52.45%) |
|  | ii. Female | 87 (47.5%) |
| iii. | **Comorbidities** |  |
|  | Jarco-Levin syndrome | 1 (0.54%) |
|  | ASD with pulmonary hypertension | 1 (0.54%) |
|  | Epilepsy | 2 (1.09%) |
|  | Down syndrome | 3 (1.63%) |
|  | Nephrocalcinosis | 1 (0.54%) |
|  | Seizure Disorder with global delay | 1 (0.54%) |
|  | Complex congenital heart disease with situs inversus | 1 (0.54%), |
|  | Splenomegaly | 1 (0.54%) |
|  | Syndromic developmental delay | 2 (1.09%), |
|  | Cerebral palsy | 4 (2.18%) |
|  | Hypo calcemic fits | 1 (0.54%) |
|  | Storage disorder | 1 (0.54%) |
|  | Congenital diaphragmatic hernia febrile fits | 1 (0.54%) |
|  | Anemia | 180 (98%) |
|  | Hereditary spherocytosis | 1 (0.54%) |
|  | Enteric fever | 1 (0.54%) |
|  | Pancytopenia | 1 (0.54%) |
|  | Meningitis | 1 (1.09%) |
|  | GERD aspiration pneumonia | 1 (0.54%) |
| iv. | **Outcomes** |  |
|  | i. Discharged | 177 (96.72%) |
|  | ii. Expired | 4 (2.18%) |
|  | iii. LAMA | 2 (1.09%) |
| v. | **Vaccination status** |  |
|  | i. Fully vaccinated | 49 (26.77%) |
|  | ii. Partially vaccinated. | 53 (28.96%) |
|  | iii. Not vaccinated | 81 (44.26%) |
| vi. | **Antiviral therapy** |  |
|  | i. Given | 110 (60.10%) |
|  | ii. Not given | 73 (39.8%) |
| vii. | **Average duration of hospitalization** | 5.74 0.2 days |
| **Association of hospital stay** |  |  |
| **Groups** |  | **P value** |
| **i.** | Infants | 0.000 |
| **ii.** | Toddlers | 0.000 |
| **iii.** | Infant-Toddlers | 0.953 |
| **Association of Symptoms** |  |  |
| **Pneumonia** |  |  |
| **i.** | Infants - Toddlers | 0.025 |
| **ii.** | Infants - Middle childhood | 0.000 |
| **iii.** | Toddlers - Middle childhood | 0.015 |
| **iv.** | Early childhood- Middle childhood | 0.016 |
| **Encephalitis** |  |  |
| **i.** | Infants - Toddlers | 0.005 |
| **ii.** | Infants – Early childhood | 0.008 |

\***ASD:** Atrial Septal Defect, **GERD:** Gastroesophageal Reflux Disease, **LAMA:** Left Admission without Medical Advice.

**Table 2.** Details of amino acid variations and their properties in query sequences and reference sequences.

| Sr no. | Amino acid position | Reference seq. WLY62623.1 | Charge /polarity | Query sequences |  |  |  |  |
| --- | --- | --- | --- | --- | --- | --- | --- | --- |
|  |  |  |  | Charge/polarity | QAUPAK1 | QAUPAK2 | QAUPAK3 | QAUPAK4 |
| 1. | 394 | Valine | Uncharged/non polar | -ve charged/polar | Aspartic acid | -- | -- | -- |
| 2. | 422 | Serine | Uncharged/polar | Uncharged/nonpolar | Glycine | Glycine | Glycine | -- |
| 3. | 487 | Serine | Uncharged/polar | +ve charged/polar | -- | -- | -- | Arginine |

**Fig 3.**
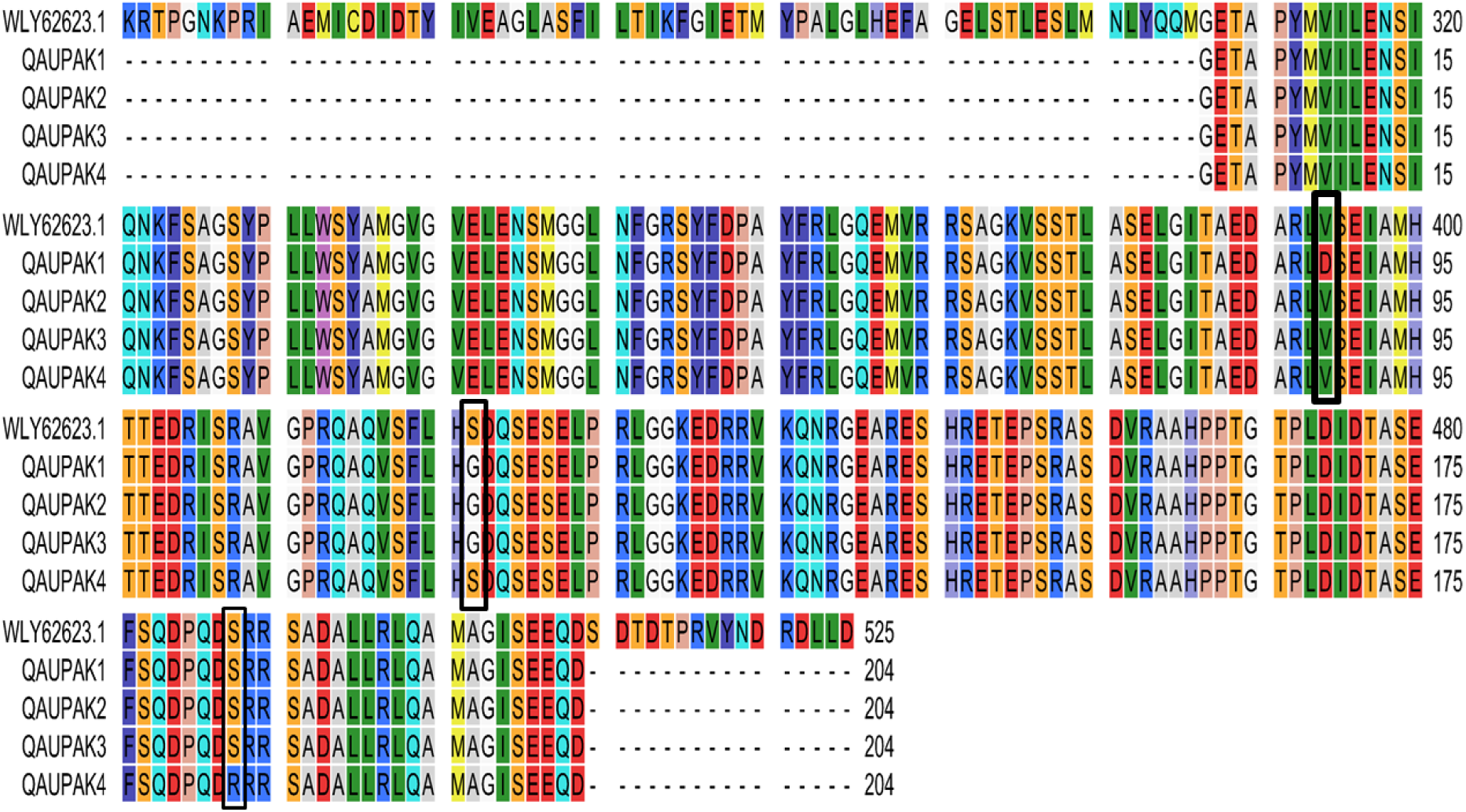
Amino acid sequence information of query sequences (QAUPAK1, QAUPAK2, QAUPAK3, QAUPAK4) w.r.t., reference sequence (WLY62623.1)

### 3.8. Secondary and Tertiary Structures Prediction

Online server PSIPRED was utilized for the investigation of secondary structures of all the isolates and reference proteins. The server provided information about the Beta strand, Alpha Helix, and Extended Coils as shown in Figure 4. The results illustrated that there is almost no difference present in the secondary structure. The percentage of Beta strands, Alpha Helix, and Extended Coils are given in Table 3. Moreover, the Robetta server was utilized for the tertiary structure of isolates and reference proteins. The tertiary structures of our isolates were compared with reference protein and amino acid substitutions were identified using Phymol. The 3D structure of isolates and reference protein along with amino acid substitutions are given in Figure 5.

**Fig 4.**
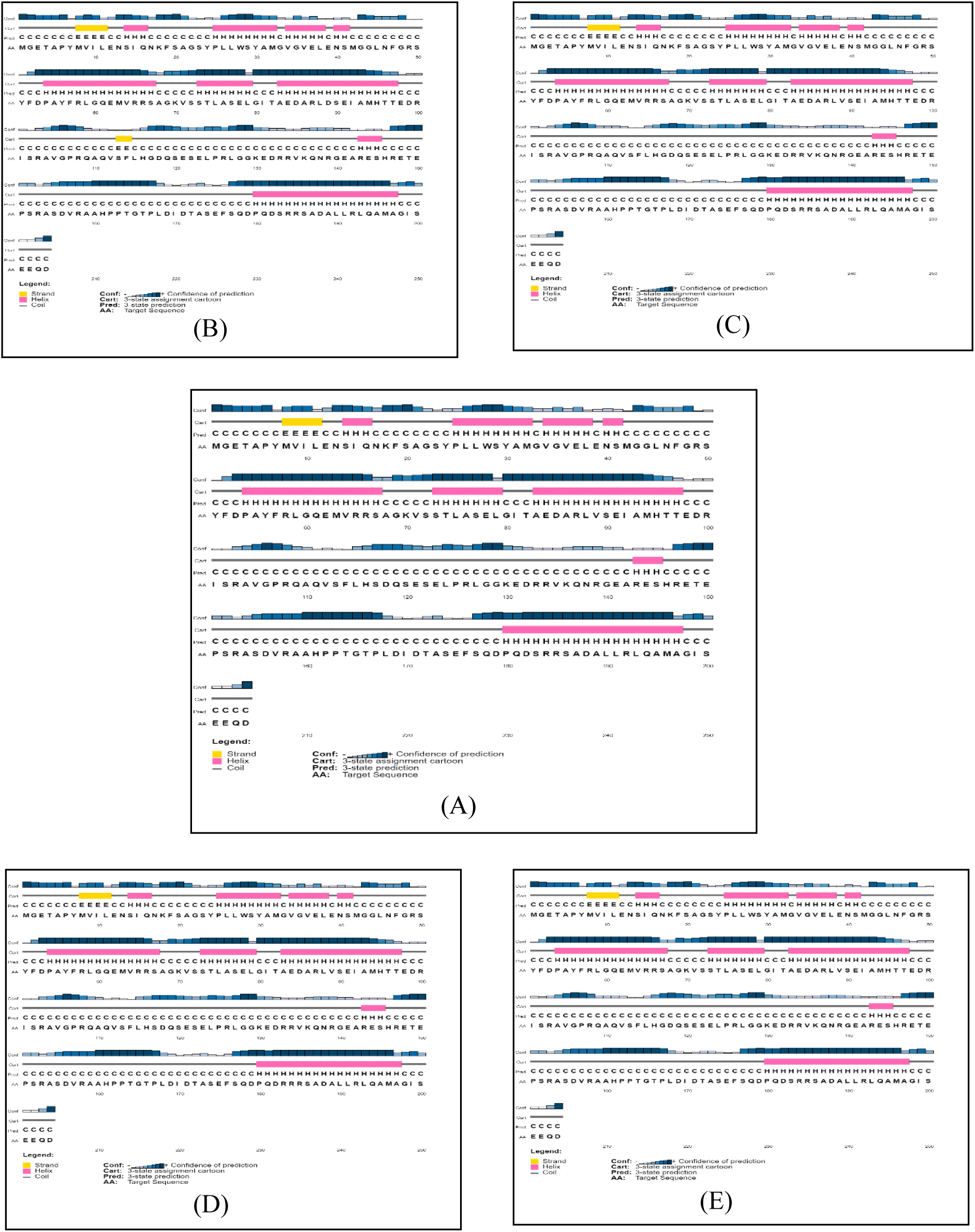
Secondary structure representation of all isolates and reference protein. (A), (B), (C), (D) and (E) represents reference protein, QAUPAK1, QAUPAK2, QAUPAK3 and QAUPAK4 respectively. Yellow color shows Beta strands, Pink color shows Alpha Helix while Blue color shows Extended Coils

**Table 3.** Percentage of Beta strand, Alpha helix, and Extended coil in reference protein and all isolates

| Sequences | Beta strand | Alpha helix | Extended coil |
| --- | --- | --- | --- |
| WLY62623.1 | 1.96% | 36.76% | 61.27% |
| QAUPAK1 | 2.94% | 36.76% | 60.29% |
| QAUPAK2 | 1.96% | 36.76% | 61.27% |
| QAUPAK3 | 1.96% | 36.76% | 61.27% |
| QAUPAK4 | 1.96% | 36.76% | 61.27% |

**Fig 5.**
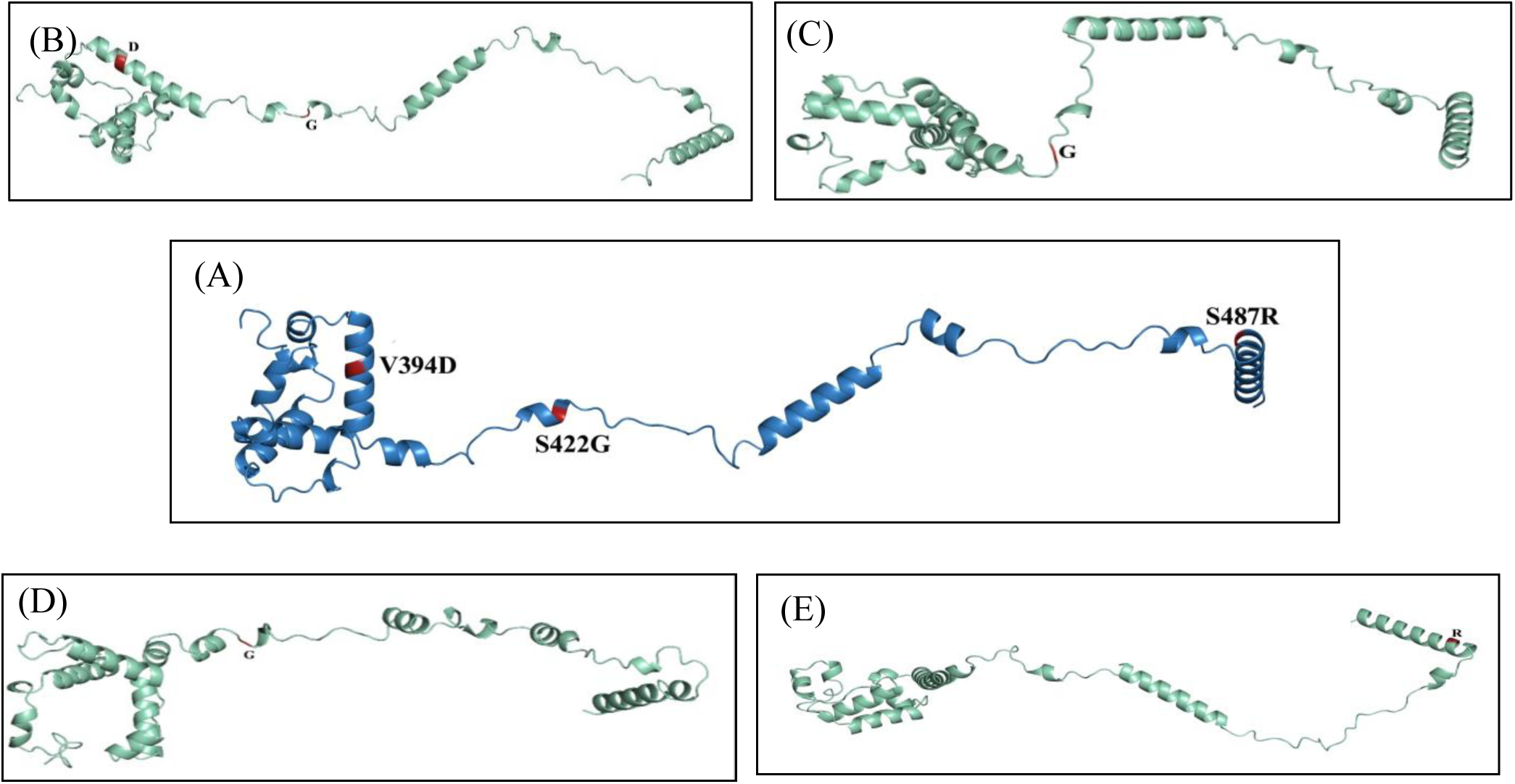
Tertiary structure representation of all isolates and reference protein. (A) Represents reference protein (Blue color) with amino acid substitution (Red color). (B), (C), (D), and (E) represent QAUPAK1, QAUPAK2, QAUPAK3, and QAUPAK4 respectively (Cyan green color) with amino acid substitutions (Red color)

## 4. Discussion

From November 2022 to April 2023, 6426 new measles cases were reported in Pakistan [17]. Measles usually gives rise to many different complications that increase inpatient hospitalization [18]. Our study explored illness severity and viral genetic characterization among hospitalized children during the 2022-2023 epidemic in Pakistan. It revealed a relatively high incidence (73.77%) of measles among infants (<12 months) and a greater number of male patients (52.45%) than females (47.5%) out of a total of 183 patients included in the present study. Moreover, 71.85% patients among infants’ group were below 9 months. Measles is particularly common among children of younger age, but it is the first time that a significant ratio of measles patients younger than 9 months have been witnessed unlike pre - Covid-19 measles outbreaks in Pakistan [1] A variety of comorbidities related to neurological, cardiac, cognitive, and pulmonary disorders were reported among hospitalized patients and there was an average of 5.74 days duration for hospitalization with the majority being discharged (96.72%) and a 2.18% mortality rate. The first dose of measles vaccine is administered at 9^th^ month and second is recommended during the second year of life for children in Pakistan [19]. The mean age for the measles patients in our study population was 16.75 months, still a great fraction (44.26%) of children had not received even their 1^st^ dose of measles vaccine which emphasizes over the need of optimization of vaccination coverage. The statistical tests revealed a significant association diseases severity with younger age groups including infants and toddlers irrespective of gender, which reaffirms the previous studies where younger age groups have been affected more by measles than older ones [20]. Pneumonia and encephalitis showed significant association with toddlers and infant age groups respectively, just like various past studies where the high incidence of such complications among measles patients has frequently been reported [21–23].

The phylogenetic analysis revealed that the isolates belong to the B3 genotype and were evolutionarily close to the viral isolates reported from the Russia and USA. Recent studies have identified the presence of B3 genotype in measles cases reported from both USA and European regions [24–25], implying genetic affinity and potential cross border transmission into Pakistan. Our results are consistent with the findings from past studies where the B3 genotype is the most prevalent worldwide and has been detected in the Pakistani region in various outbreaks during the past two decades [1, 26]. However, a few cases from other genotypes including H1 and D genotypes had also been reported in some regions during previous outbreaks [26].

Nucleoprotein is the most abundant, highly immunogenic structural protein that plays a crucial role in the regulation of replication, transcription, genome packaging and gene expression of morbilliviruses. [27–29]. It comprises 525 amino acids and contains two domains i.e. the N_CORE_ (residues 1-400) interacts with the viral RNA and maintains nucleocapsid structure and the N_TAIL_ (residues 401-525) that is a long intrinsically disordered domain that anchors the polymerase complex. Disordered N_TAIL_ contains a molecular recognition element (MoRE) (residues 485-502) whose interaction with the C-terminal three-helix bundle domain XD of phosphoprotein (residues 459-507) leads to the recruitment of polymerase complex onto the nucleocapsid template [30]. Our study encompasses the analysis of 203 C-terminal residues (306-509) that form the anterior portion of the N_CORE_ domain and a major (87.2%) portion of the N_TAIL_ domain including molecular recognition element (MoRE). The substitutional analysis of 203 amino acids from the C-terminal of nucleoprotein showed a total of 3 substitutions concerning the reference sequence (WLY62623.1). In QAUPAK1 the V394D substitution was recorded in the terminal portion of N_CORE_ domain. Valine is a medium-sized, hydrophobic, aliphatic, uncharged polar amino acid that prefers to be buried in protein hydrophobic cores. Valine is Cβ branched which means that unlike most of the other amino acids, they have two non-hydrogen substituents attached to its Cβ carbon which means that there is more bulkiness around the protein backbone and more restrictions in the conformation that the main chain can assume as a result valine is least likely to assume α helical structure. Valine has been substituted by Aspartic acid, which is a small, hydrophilic, negatively charged polar amino acid, and contrary to valine it prefers to be on the surface of protein [31]. Aspartic acid is frequently involved in the formation of salt bridges and its negative charge confers the ability to interact with the positively charged non-protein atoms such as zinc hence they are frequently involved in the protein active or binding sites. We can speculate that substitution with amino acids having opposite attributes can have a possible effect on the functioning of the N_CORE_ domain and the possibility of altered interaction with the viral RNA and maintenance of nucleocapsid structure [32]. The second substitution S422G was seen uniformly across QAUPAK1, QAUPAK2, and QAUPAK3. Both serine and glycine are very small uncharged amino acids with the former being polar and later nonpolar. Serine is distinct from other amino acids with hydrogen as a side chain rather than carbon like all other amino acids. With this uniqueness in structure if a conserved serine changes into any other amino acid there can be a significant functional impact. Another substitution S487R was recorded that lies in the N_TAIL_ region where serine was replaced by arginine which is a positively charged polar amino acid. Arginine mostly forms part of the active site and interacts with negatively charged proteins [31]. Due to diverse roles of N protein in Measles virus life cycle, we can speculate that the substations in the amino acid sequence of N gene would have impact on the structure and function of Measles virus [29].

The secondary structures were built *de novo* for viral isolates (QAUPAK1, QAUPAK2, QAUPAK3, QAUPAK4), and reference sequence (WLY62623) which showed a greater degree of similarity with some minor changes in the orientation of protein 3D structure. All the isolates formed a higher ratio of ≥ 60 of extended coil structure, 36.76% of α helix, and >1% of β strand. The variations in the protein structure can be due to the intrinsic disorders in the nucleoprotein of morbilliviruses or may be due to the amino acid substations observed in the study.

Measles in Pakistan remains a persistent challenge due to several contributing factors. While children younger than 9 months typically acquire immunity from maternal antibodies against measles, the high prevalence of the disease in this age group is alarming. Our study did not record the maternal medical or vaccination history, suggesting a need for future research focusing on women of childbearing age. Investigating post-COVID changes in maternal immunity could provide valuable insights into the prevalence of measles in young children. Furthermore, children are often admitted to hospitals at later stages of the disease when the viral load is declining, complicating molecular detection efforts. The simultaneous admission of multiple children from the same family underscores the necessity of raising awareness about measles infectiousness. Promoting early admission of measles patients could help prevent the spread of infection to healthy children. Additionally, our study focused on a single gene that exhibited significant substitutions and structural changes. Future studies should extend to other critical genes, exploring their mutational, structural, and functional characteristics. This could offer deeper insights into potential viral mutations that may be compromising the efficacy of current vaccines and treatments.

## 5. Conclusion

In conclusion, our study highlights the critical importance of integrating epidemiological and molecular approaches to explain the complexities of measles outbreaks, particularly among the concerning surge observed globally and especially in Pakistan between 2022 and 2023. By meticulously analyzing both the epidemiological data and molecular characteristics of the measles virus (MV), we have elucidated the severity of the disease among hospitalized children, the persistent circulation of the B3 genotype, and the significant variations in viral structure and amino acid substitutions, potentially influencing treatment effectiveness. Our findings emphasize the necessity for continuous surveillance, complete gene sequencing, and a deeper understanding of strain variations at both structural and functional levels to inform more efficacious treatment strategies and alleviate the burden on healthcare systems. This study serves as a crucial step towards combating measles outbreaks and offers valuable insights applicable to the management of future infectious disease challenges.

## Data Availability

The data will be available from the corresponding author upon reasonable request.

## Acknowledgements

We acknowledge the admitted patients, their guardians, and the PIMS staff for their cooperation during the sample collection.

## Author’s contribution

AN, SR and MA conceptualized the study; AN and NH samples collection and data collection; AN, AZ, and SR conducted data analysis and interpretation. AN and SR wrote the first draft of the manuscript, and all authors provided input on the various versions of the draft manuscript and reviewed and approved the final manuscript. MA supervised the research.

## Ethics Approval and Consent to Participate

This study was approved by the Bio-Ethical Committee (BEC-FBS-QAU2023-520) at Quaid-i-Azam University Islamabad. Participants provided informed consent to participate in the study.

## Availability of Data and Material

The data will be available from the corresponding author upon reasonable request.

Permission to reproduce material from other sources:

In this study, we didn’t reproduce material from other sources.

